# Recent Trends in Aspirin Use for Cardiovascular Disease Prevention in the United States - 2015-2023

**DOI:** 10.1101/2022.08.22.22278874

**Authors:** Karthik Murugiah, Claudia See, Chenxi Huang

## Abstract

**BACKGROUND:** Recent trials show a lack of benefit of routine aspirin use for primary prevention. We assessed recent trends in aspirin use for both primary and secondary prevention in the United States population.

**METHODS:** We used National Health Interview Survey (NHIS) data from 2015-2023 and reported the annual weighted proportion of aspirin use among participants ≥40 years without a self-reported history of angina, coronary heart disease, myocardial infarction, or stroke (primary prevention cohort) and those with (secondary prevention cohort), overall and by subgroups. Statistical comparisons were made only between 2019 and 2023 due to an NHIS study redesign in 2019.

**RESULTS:** From 2019 to 2023 overall aspirin use for primary prevention decreased from 20.6% to 15.7%. It declined in all subgroups, including among individuals ≥70 years from 38.7% to 30.7%, and those age <60 years but with ≥3 CVD risk factors (29.1% to 19.7%). Overall aspirin use for secondary prevention also declined, but to a lesser extent from 65.7% to 61.9%. (all P <0.001)

**CONCLUSION:** In response to recent data there has been a significant reduction in aspirin use for primary prevention, appropriately among older participants, but also among younger participants with multiple cardiovascular risk factors. There was also a minor decrease in its use for secondary prevention which warrants further study.

## INTRODUCTION

Aspirin has long been used to reduce cardiovascular events among individuals at increased atherosclerotic cardiovascular disease (ASCVD) risk. However, recent trials in 2018 question routine use of aspirin for primary prevention and suggest potential harm in older adults (1-4). In response, in 2019, the American Heart Association and American College of Cardiology (AHA/ACC) updated their guidelines, discouraging routine aspirin in persons > 70 years and those with increased bleeding risk (5). In 2022, the U.S. Preventive Services Task Force (USPSTF) recommended against aspirin for primary prevention among adults ≥60 years.

This recommendation change represents a paradigm shift in cardiovascular prevention. A recent study using the National Health Interview Survey (NHIS) suggested that aspirin use for primary prevention is declining.(6) However, the study was limited to 2021 and it is unknown if these declines have continued further, especially in light of the 2022 USPSTF recommendations.(7) Additionally, it is unknown if the decline in aspirin use varies by baseline cardiovascular risk factors. Further, due to an error by NHIS, questions about aspirin use were not fielded in Quarter 1 of 2021 and thus the estimates of aspirin use in 2021 may not be nationally representative.

Accordingly, we studied trends in aspirin use for primary and secondary prevention using 2015-2023 data from the NHIS - a principal source of information on civilian noninstitutionalized US population health (8). We reported aspirin use overall and by subgroups of age (40-59, 60-69 and ≥70 years), sex, and diabetes. In the primary prevention subgroup, aspirin use was additionally assessed among those <60 years but with ≥3 traditional ASCVD risk factors (hypertension, hypercholesterolemia, obesity, diabetes, and current smoking).

## METHODS

Data from the NHIS Sample Adult files 2015-2023 were used. Aspirin use questions were changed to a biennial schedule in 2019 and thus aspirin use in 2020 and 2022 is not reported.

Primary and secondary prevention cohorts included participants ≥40 years without and with ASCVD (self-reported history of angina, coronary heart disease, myocardial infarction, or stroke), respectively. Participants reporting taking aspirin on medical advice or self-use were classified as aspirin users.

All analyses incorporated sampling weights to produce nationally representative estimates. Aspirin use was compared using Chi squared tests only between 2019 and 2023 due to a redesign of the NHIS sampling weights calculation in 2019 to address non-response bias, which may affect comparisons between 2019 and earlier years.

Analyses were conducted using Stata v17, College Station, TX. Significance level was set at a two-sided p-value of 0.05. As NHIS data are public without identifiers, this study was exempt from Yale Institutional Review Board review.

## RESULTS

Our study sample included 139,400 participants over the entire study period representing approximately 149 million adults ≥40 years annually. From 2019 to 2023, overall aspirin use for primary prevention decreased from 20.6% to 15.7%. It declined in all subgroups, including among individuals ≥70 years from 38.7% to 30.7% and those age <60 years but with ≥3 CVD risk factors (29.1% to 19.7%). Overall aspirin use for secondary prevention also declined, but to a lesser extent from 65.7% to 61.9%. (all P<0.001) (Figure 1).

**Figure 1:**
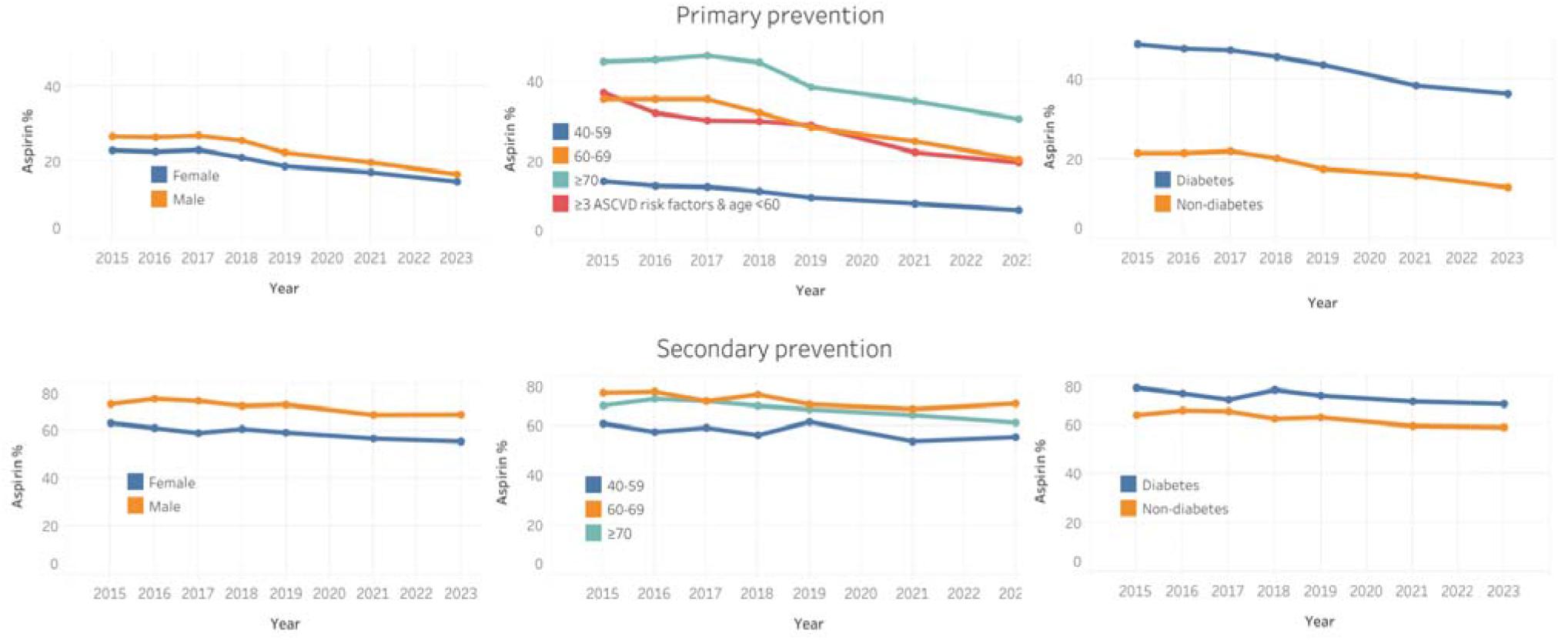
Weighted Proportion of Individuals on Aspirin for Primary and Secondary Prevention by subgroups All Chi-squared P for comparison 2019 vs 2023 <0.001 ASCVD: Atherosclerotic Cardiovascular Disease

## DISCUSSION

Our study shows the impact of recent aspirin trial data and guideline changes on aspirin use on a nationally representative US population sample. As expected, aspirin use for primary prevention declined significantly. Among individuals ≥ 70 years - a population unlikely to benefit from aspirin for primary prevention given bleeding risk (4), an 8% absolute and 18% relative decline in aspirin use occurred between 2019 and 2023. This reduction matches the Class III recommendation for aspirin as primary prevention in this age group from 2019 ACC/AHA guidelines (5). However, despite this reduction, a third of adults ≥ 70 continued to take aspirin for primary prevention with a net potential for harm. On the contrary, a 25% relative decline in aspirin use also occurred among individuals <60 years and with ≥ 3 traditional ASCVD risk factors, who may be at higher than average risk, and may benefit from aspirin (5,7). A similar degree of decline in aspirin use in both older adults and younger adults at elevated ASCVD risk is puzzling and suggests that some aspirin discontinuations may be occurring without discussion with physicians.

Aspirin use for secondary prevention also decreased, especially in the younger age group, but to a lesser degree than among primary prevention users. This trend may be partly due to physicians prescribing single P2Y12 inhibitor agents instead of aspirin for secondary prevention, a practice increasingly supported by data (10). Also, physicians may be avoiding additional aspirin in patients with other indications for chronic anticoagulation. Future studies are needed to study this reduction in aspirin use for secondary prevention in the context of P2Y12 and anticoagulation therapy. Concern remains that some patients may have misinterpreted media messaging on aspirin and stopped taking aspirin prescribed for secondary prevention.

This study has its limitations. First, the NHIS lacks questions on peripheral arterial and atherosclerotic aortic disease which may benefit from aspirin, or colorectal cancer risk for which aspirin has been suggested. Similarly, we lack data on hemorrhagic stroke history and individual bleeding risk. Second, we have no blood pressure and cholesterol measurements to assess ASCVD risk. Third, adults younger than 40 years were not queried regarding aspirin use in the NHIS survey. Fourth, the 2019 NHIS redesign may affect comparisons with previous years. Finally, the NHIS relies on self-reported aspirin use which may be affected by recall bias.

## CONCLUSION

In response to recent data, aspirin use has decreased for primary prevention, both among older adults which is appropriate, but also younger adults at elevated risk which may be cause for concern. Aspirin use for secondary prevention also reduced which needs further study. Physicians must ensure their patients are informed and on appropriate prevention regimens.

## Data Availability

All data produced are available online at https://www.cdc.gov/nchs/nhis/data-questionnaires-documentation.htm

## REFERENCES

1. Gaziano JM, Brotons C, Coppolecchia R et al. Use of aspirin to reduce risk of initial vascular events in patients at moderate risk of cardiovascular disease (ARRIVE): a randomised, double-blind, placebo-controlled trial. The Lancet 2018;392:1036–1046.

2. McNeil JJ, Nelson MR, Woods RL et al. Effect of Aspirin on All-Cause Mortality in the Healthy Elderly. New England Journal of Medicine 2018;379:1519–1528.

3. Effects of Aspirin for Primary Prevention in Persons with Diabetes Mellitus. New England Journal of Medicine 2018;379:1529–1539.

4. McNeil JJ, Wolfe R, Woods RL et al. Effect of Aspirin on Cardiovascular Events and Bleeding in the Healthy Elderly. New England Journal of Medicine 2018;379:1509–1518.

5. Arnett DK, Blumenthal RS, Albert MA et al. 2019 ACC/AHA Guideline on the Primary Prevention of Cardiovascular Disease: A Report of the American College of Cardiology/American Heart Association Task Force on Clinical Practice Guidelines. Circulation 2019;140:e596–e646.

6. Gupta M, Gulati S, Nasir K, Sarraju A. Aspirin Use Prevalence for Cardiovascular Disease Prevention Among US Adults From 2012 to 2021. Annals of Internal Medicine 2024.

7. Davidson KW, Barry MJ, Mangione CM et al. Aspirin Use to Prevent Cardiovascular Disease: US Preventive Services Task Force Recommendation Statement. Jama 2022;327:1577–1584.

8. Centers for Disease Control and Prevention. About the National Health Interview Survey. Reviewed March 3, 2022. Accessed August 21, 2022.

9. Potential Impact of NHIS Redesign and COVID-19 on the Cancer Trends Progress Report.

10. Chiarito M, Sanz-Sánchez J, Cannata F et al. Monotherapy with a P2Y12 inhibitor or aspirin for secondary prevention in patients with established atherosclerosis: a systematic review and meta-analysis. The Lancet 2020;395:1487–1495.

